# Examining real-world Alzheimer’s disease heterogeneity using neuroanatomical normative modelling

**DOI:** 10.1101/2022.11.02.22281597

**Authors:** Flavia Loreto, Serena Verdi, Seyed Mostafa Kia, Aleksandar Duvnjak, Haneen Hakeem, Anna Fitzgerald, Neva Patel, Johan Lilja, Zarni Win, Richard Perry, Andre F. Marquand, James H. Cole, Paresh Malhotra

## Abstract

Alzheimer’s disease (AD) has been traditionally associated with episodic memory impairment and medial temporal lobe atrophy. However, recent literature has highlighted the existence of atypical forms of AD, presenting with different cognitive and radiological profiles. Failure to appreciate the heterogeneity of AD in the past has led to misdiagnoses, diagnostic delays, clinical trial failures and risks limiting our understanding of the disease. AD research requires the incorporation of new analytic methods that are as free as possible from the intragroup homogeneity assumption underlying case-control approaches according to which patients belonging to the same group are comparable to each other. Neuroanatomical normative modelling is a promising technique allowing for modelling the variation in neuroimaging profiles and then assessing individual deviations from the respective distribution. Here, neuroanatomical normative modelling was applied for the first time to a real-world *clinical* cohort of Alzheimer’s disease patients (n=86) who had a positive amyloid PET scan and a T1-weighted MR performed as part of their diagnostic workup. The model indexed normal cortical thickness distributions using a separate healthy reference dataset (n= 33,072), employing hierarchical Bayesian regression to predict cortical thickness per region using age and sex. Transfer learning was used to recalibrate the normative model on a *validation cohort* (n=20) of scanner-matched cognitively normal individuals. Brain heterogeneity was quantified as *z-scores* at each of the 148 ROIs generated within each AD patient. Z-scores < -1.96 defined as outliers. Clinical features including disease severity, presenting phenotypes and comorbidities were collected from health records to explore their association with outlier profiles. Amyloid quantification was performed using an automated PET-only driven method to examine the association between amyloid burden and outliers.

The total number of individual outliers (*total outlier count*) in biomarker-confirmed AD clinical patients ranged between 1 and 120 out of 148 (median 21.5). The superior temporal sulcus was the region with the highest count of outliers (60%) in AD patients. The mean proportion of outliers was higher in the temporal (31.5%) than in the extratemporal (19.1%) regions and up to 20% of patients had no temporal outliers. We found higher mean outlier count in patients with non-amnestic phenotypes, at more advanced disease stages and without depressive symptoms. Amyloid burden was negatively associated with outlier count. This study corroborates the heterogeneity of brain atrophy in AD and provides evidence that this approach can be used to explore anatomo-clinical correlations at an individual level.

## Introduction

For decades, the ‘typical’ Alzheimer’s disease (AD) patient has been described as an older adult with marked episodic memory impairment and loss of grey matter volume in the medial temporal lobe (MTL). However, over recent years, it has become increasingly clear that AD is more heterogeneous than previously thought and can present with several forms which vary in age of onset, clinical presentation, neuropathological and genetic profiles ^1^. Moreover, research into its earlier phases has highlighted that AD is a continuum, rather than a discrete clinical entity, that goes from normal cognitive status to mild cognitive impairment to dementia ^2 3^. Moreover, AD often coexists with other neuropathologies ^4^ or clinical comorbidities ^5-8^ that contribute to interindividual heterogeneity. In a recent study evaluating a memory clinic cohort, we have found that about half of patients requiring biomarker confirmation of AD due to diagnostic uncertainty also reported depressive symptoms ^8^. Hypertension, heart disease and diabetes are also commonly comorbid with AD.

Advances in diagnosis, treatment and understanding of the pathophysiological mechanisms of AD require research to move beyond the idea of a ‘typical’ AD patient ^9^ as this implies an interindividual similarity that is not reflected in the real-world clinical population. Not all AD patients present with a typical phenotype and age of onset, and failure to recognise this frequently leads to diagnostic delays and errors ^10^. Efforts to incorporate the concept of AD as a continuum and as a heterogeneous condition into the clinical framework are illustrated by the evolution of AD diagnostic guidelines, which have been giving increasing weight to biological markers of AD to diagnose it accurately and at its earlier stages ^11, 12^. On the other hand, AD research has yet to progress towards analysis methods that make interindividual differences the focus rather than a source of error. Finding ways to dissect the heterogeneity of clinical and neuropathological profiles has become one of the central priorities in this field ^13^.

The dominant approach in case-control studies is to compare the average patterns of atrophy of AD patients with those of healthy individuals. While this method has been fundamental for the detection of important hallmarks of ‘typical’ AD, such as MTL atrophy ^14^, it provides limited information about disease mechanisms across individuals ^15^. AD patients are grouped together, hence considered comparable to each other and clearly distinct from healthy controls. This implies assumption of intragroup homogeneity and conceives the disease as a discrete entity rather than as a spectrum. Moreover, this approach carries the notion that the typical population likely represents a more homogeneous group that is presumably a more adaptive or optimal state ^16^, making it the reference standard for AD research and clinical trials. This is in contrast with recent studies that have demonstrated intragroup variability even within the clinical population of ‘typical AD’ ^17^. Making heterogeneity the object of study requires to move beyond the classic case-control approach and use techniques that address interindividual variation.

As neuroimaging has entered the era of big data, data-driven clustering methods have been the predominant avenue for the exploration of heterogeneity in dementia (see Habes et al., 2020 ^18^ for a review). These have led to the discovery of neuropathological subtypes of AD associated with different distribution of tau neuropathology ^19, 20^, patterns of atrophy ^21, 22^ and of hypometabolism ^23^, and cognitive and demographic characteristics ^19, 24^, but comparable amyloid distribution ^25, 26^. Although these studies marked significant progress in the comprehension of AD heterogeneity, their generalisability requires prior evaluation of key potential issues^18 27 15, 27, 9, 28^.

Neuroanatomical normative modelling is an emerging statistical technique, alternative and complementary to clustering ^15^, which shifts the focus from group averages to cohort variation ^28^. Similar to growth charts used in paediatric medicine, this method aims to gather individual-level information by comparison with the norm ^15, 28^. This is done by estimating centiles of variation of a brain measure (e.g., cortical thickness) across a normative population and then assessing how much each individual deviates from the respective distribution ^9^. Moreover, it allows spatially mapping of the extent to which an individual deviates from the norm at any given brain region, providing a map of individual variability ^9^. This technique has been used in psychiatric research over the last years with the aim of conceptualizing mental disorders as deviations from expected functioning and parsing disease heterogeneity ^29^. Recent studies have used normative modelling to map interindividual differences in brain structure among patients with attention-deficit hyperactivity disorder ^30^, bipolar disorder ^31, 32^, autism ^33-36^, and schizophrenia ^31, 32^. On the other hand, this technique has had limited use in AD research so far ^37, 38^, despite its potential to unveil disease heterogeneity. In a recent application by Verdi and colleagues, neuroanatomical normative models revealed a largely heterogeneous distribution of cortical atrophy in a group of AD patients from the Alzheimer’s Disease Neuroimaging Initiative (ADNI) research cohort ^39^ (where the unwanted noise from acquisition across multiple sites was modelled using a hierarchical Bayesian regression^40^). Moreover, while previous studies have examined the association between amyloid and mean cortical thickness ^41^, no study to date has investigated how amyloid burden relates to individual-level deviations in cortical thickness from the norm.

In the present study, for the first time, neuroanatomical normative modelling employing hierarchical Bayesian regression is applied to a real-world clinical cohort in order to map disease variation in diagnostically challenging patients who required biomarker confirmation of Alzheimer’s disease. Patients from the Imperial Amyloid PET Cohort (APC) were included in this study if they had an MRI and an amyloid PET as part of their diagnostic investigations. This clinical cohort was established at Imperial College Healthcare NHS Trust in 2013 and includes all patients seen at the Imperial Memory Clinic and receiving amyloid PET imaging as part of their diagnostic workup ^42^. The objectives of this study were to: (i) assess intragroup neuroanatomical heterogeneity in a real-world clinical cohort of patients with confirmed AD; (ii) evaluate the use of normative modelling to explore anatomo-clinical correlations at an individual level; (iii) examine the association between global amyloid burden and deviations in cortical thickness.

## Material and Methods

### Subjects

We reviewed 256 amyloid-positive patients from the Imperial APC Cohort who had a clinical amyloid PET scan between 2014 and 2021 as part of their diagnostic workup. At Imperial College Healthcare NHS Trust (ICHT), all amyloid PET referrals are made by consensus within a multidisciplinary team ^42^ and are in line with the Amyloid Imaging Taskforce’s appropriate use criteria ^43^. Patients were included in this study if they had a positive amyloid PET and an available clinical MRI scan performed within 12 months of amyloid PET (n=186). Of these, 80 had to be excluded following preliminary inspection of MRI images due to unavailable or ineligible T1-weighted images, motion artefacts, other pathologies affecting brain integrity (i.e., normal pressure hydrocephalus, multiple sclerosis, large infarcts) or previous neurosurgery, leaving a total of 106 subjects, hereafter termed the ‘*clinical cohort’*. As explained in greater detail below, the normative modelling framework also requires a reference and a validation cohort to respectively build and validate the model. The ‘*reference cohort’* consisted of a large group of 33,072 cognitively normal adults pooled from publicly available neuroimaging datasets under a previous study ^39^; the relevant methods are described elsewhere ^39^. The ‘*validation cohort’* constituted a group of 28 cognitively normal (CN) older adults who had an MRI scan for research purposes at ICHT.

### MR image acquisition

All subjects had whole-brain T1-weighted volumetric images. A total of 86 (81%) patients from the clinical cohort were scanned at ICHT using a 1.5T Siemens MAGNETOM Avanto with the following parameters: repetition time = 900ms; echo time = 3.37ms; 160 slices/slab, yielding voxel size of 1×0.5×0.5mm. Patients scanned at other sites were excluded from the model. All CN individuals were scanned at ICHT using a 3T Siemens MAGNETOM Verio. Image parameters were as follows: repetition time = 900ms; echo time = 2.52ms; 176 slices/slab, yielding voxel size of 1×1×1mm. The imaging protocol for the reference cohort varied across different studies ^39^.

### MR image analysis

#### Cortical segmentation

After a preliminary inspection of raw T1-weighted images, cortical reconstruction and volumetric segmentation were performed using FreeSurfer 6.0 *recon-all* function (https://surfer.nmr.mgh.harvard.edu/) ^44^. To be consistent with the reference cohort, we used the Destrieux atlas of 148 cortical parcellations (74 in each hemisphere) that are classified as gyral or sulcal ^45^. In the ‘clinical’ and ‘validation’ cohorts, the output was visually checked for quality control by three authors independently (F.L., A.D., H.H.) following preliminary training on a subset of cases to ensure consistency. Unclear issues were discussed and addressed jointly by the team. Output errors were found in 87 (82%) patients and 19 (68%) CN individuals and addressed in line with FreeSurfer guidelines (https://surfer.nmr.mgh.harvard.edu/fswiki/FsTutorial/TroubleshootingData) (examples in **Supplementary Fig.1**). After correction, the *recon-all* command was re-run to re-generate surfaces and the output underwent further visual inspection. At this stage, 2 patients had to be excluded due to persistent segmentation errors after multiple correction attempts, leaving a total of 104 patients. Cortical segmentation procedures for the *reference cohort* are described elsewhere ^39^. We extracted the estimates of total intracranial volume and cortical thickness across 148 regions of interest from the Destrieux atlas ^45^.

#### Neuroanatomical normative modelling

In this study, we used the Hierarchical Bayesian Regression proposed by Kia and colleagues which provides a solution for normative modelling on real-world clinical data because first, it handles artefactual variability in data associated with site effects; and second, due to its federated nature and model transferability, it provides the possibility for normative modelling on distributed data without the need to retrain a reference cohort^40, 46^. Hierarchical Bayesian Regression was previously trained on a *reference cohort* (compiled by Kia and colleagues), using age and sex as the covariates, to index population variability in cortical thickness across all 148 ROIs ^43^. This process modelled centiles of variations for any value of the covariates, hence describing the full range of normal variation ^15^. Transfer learning was used to recalibrate the previously run normative model to the scan site using cortical thickness data from a subgroup of scanner-matched CN participants (n=20) of the *validation cohort*. The recalibrated model was then applied to the *clinical cohort*. Deviations in cortical thickness from the normative cohort were quantified as separate *z-scores* at each ROI for each patient; z-scores < -1.96 were defined as ‘outliers’, and the count of each patient’s outlier regions was totalled (*total outlier count)*. The analysis of outliers was limited to negative deviations as the primary interest of this study was AD-related neurodegeneration as indexed by lower cortical thickness. To assess the spatial distribution of these deviations (i.e., areas with significantly lower cortical thickness), we built individualised outlier maps and computed the overlap in outlying regions across patients.

### Amyloid PET image acquisition

All clinical patients included in this study were scanned using a Siemens Biograph 64 PET/CT scanner. The PET ligand used for amyloid PET imaging changed from 18F-Florbetapir (Amyvid) to 18F-Florbetaben (Neuraceq) in December 2017, following cessation of UK (18)F-florbetapir manufacture. For 18F-Florbetapir, a 20-min acquisition of the brain was obtained following a 40-min interval post-injection of an intravenous bolus of 370 MBq (minimum 333MBq); for 18F-Florbetaben, a 30-min brain acquisition was obtained following a 90-min interval post-injection of an intravenous bolus of 360 MBq (minimum 240MBq). A low dose CT image was also acquired and used for attenuation correction and anatomical localization of the PET images ^47^. Both the Amyvid and Neuraceq images were reconstructed using OSEM 3D, 4 iterations, 14 subsets, 168 matrix size, zoom 2, Gaussian 3mm FWHM.

### Amyloid PET review and analysis

#### Clinical interpretation

As part of the diagnostic workup, all amyloid PET images underwent standard clinical interpretation: an experienced nuclear medicine radiologist classified them as ‘amyloid-positive’ or ‘amyloid-negative’ through visual reads using greyscale images and the cerebellum as the reference region ^48^. Equivocal cases were independently read by two nuclear medicine radiologists and by a third reader when there was disagreement. Amyloid PET positivity was one of the inclusion criteria of this study. A positive amyloid PET scan indicates increased burden of Alzheimer’s pathology and, in the clinical context, is highly suggestive of an AD clinical diagnosis.

#### Amyloid quantification

Quantification of amyloid PET images was performed using Hermes BRASS version 4.0 (Hermes Medical Solutions AB, Stockholm, Sweden) software package, a fully automated PET-only driven method fully described in Lilja et al. 2019 ^49^. Briefly, each PET image is first spatially normalized to Montreal Neurological Institute (MNI) space. The images are then registered to a synthetic tracer-specific adaptive template that was created using a linear combination of two tracer-specific principal component images. The two principal components were designed to span from Aβ-negativity to Aβ-positivity: the first consists in the average of all the images of the dataset, the second corresponds to the difference between Aβ-positive and Aβ-negative images ^49^. This method provides regional standardized uptake value ratio (SUV_R_) and a global Aβ index. The SUV_R_ is the ratio between tracer uptake within each ROI to that in the reference region. The reference region used in this study was the cerebellum, which has been shown to allow earlier detection of amyloid accumulation compared to the pons ^50^. The regional SUV_R_ was computed across 48 ROIs defined with a probabilistic atlas ^49^. The Aβ index is a novel measure of amyloid burden ^49, 51^ corresponding to the total weight of global amyloid deposition; its value ranges between -1 (Aβ-negative appearance) and +1 (Aβ-positive appearance). In this study, amyloid PET quantification was not used to define amyloid status, which was solely based on expert visual reads.

### Clinical measures

Alzheimer’s disease heterogeneity within the normative modelling framework can be further explored by examining how individual profiles of deviations relate to the clinical features as observed within the clinical setting. All patients included in this study had a comprehensive clinical assessment at ICHT, including medical history review, patient and caregiver interview, neurological examination, and cognitive screening. Referral letters and cognitive neurology clinical correspondence were stored within the Imperial College Healthcare NHS Trust’s electronic patient record, allowing us to retrospectively collect clinical background information relevant to this study. For all patients, we recorded whether they presented with an amnestic or non-amnestic clinical phenotype, the disease stage (mild cognitive impairment *vs* dementia stage) at the time of MRI scanning, and whether they had a history of depressive symptoms (**Table 1**). Patients were categorised as ‘amnestic’ if the predominant cognitive difficulties at presentation involved the episodic memory domain, and as ‘non-amnestic’ if disease onset was characterised by impairment in non-memory domains (namely visuospatial, behavioural or language). The classification into ‘mild cognitive impairment’ ^52^ or ‘dementia’, instead, was performed through structured review of clinical records and was in line with Petersen et al.’s criteria, which differentiate the two stages of the disease on the basis of preserved/impaired independent functioning ^53^. In addition, we systematically assessed whether patients had previous or ongoing depressive symptoms, defined as any signs of low mood that were severe enough to be discussed in the clinical notes by the referring clinician and/or dementia expert (as described in Loreto et al., 2022).

**Table 1.**
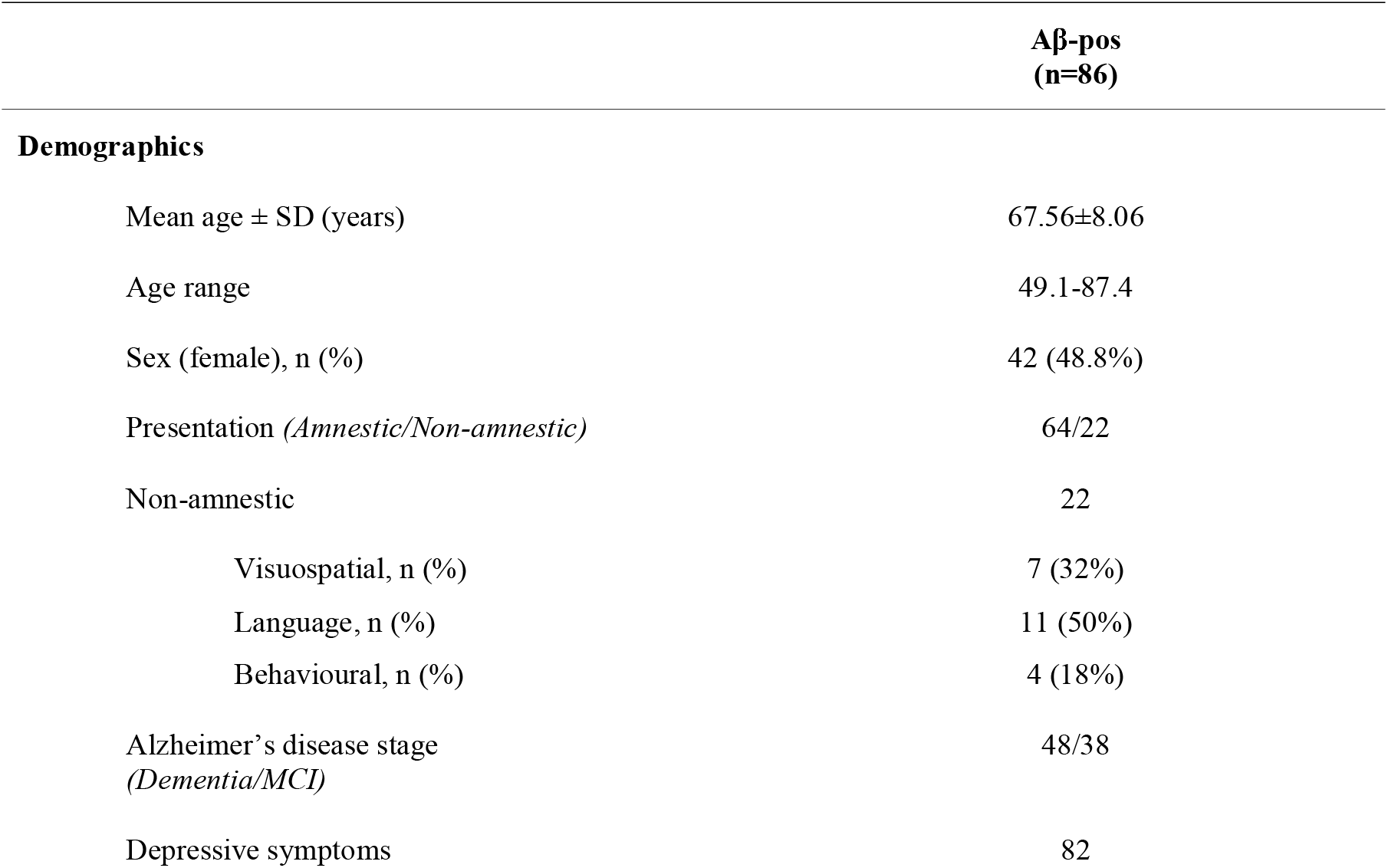
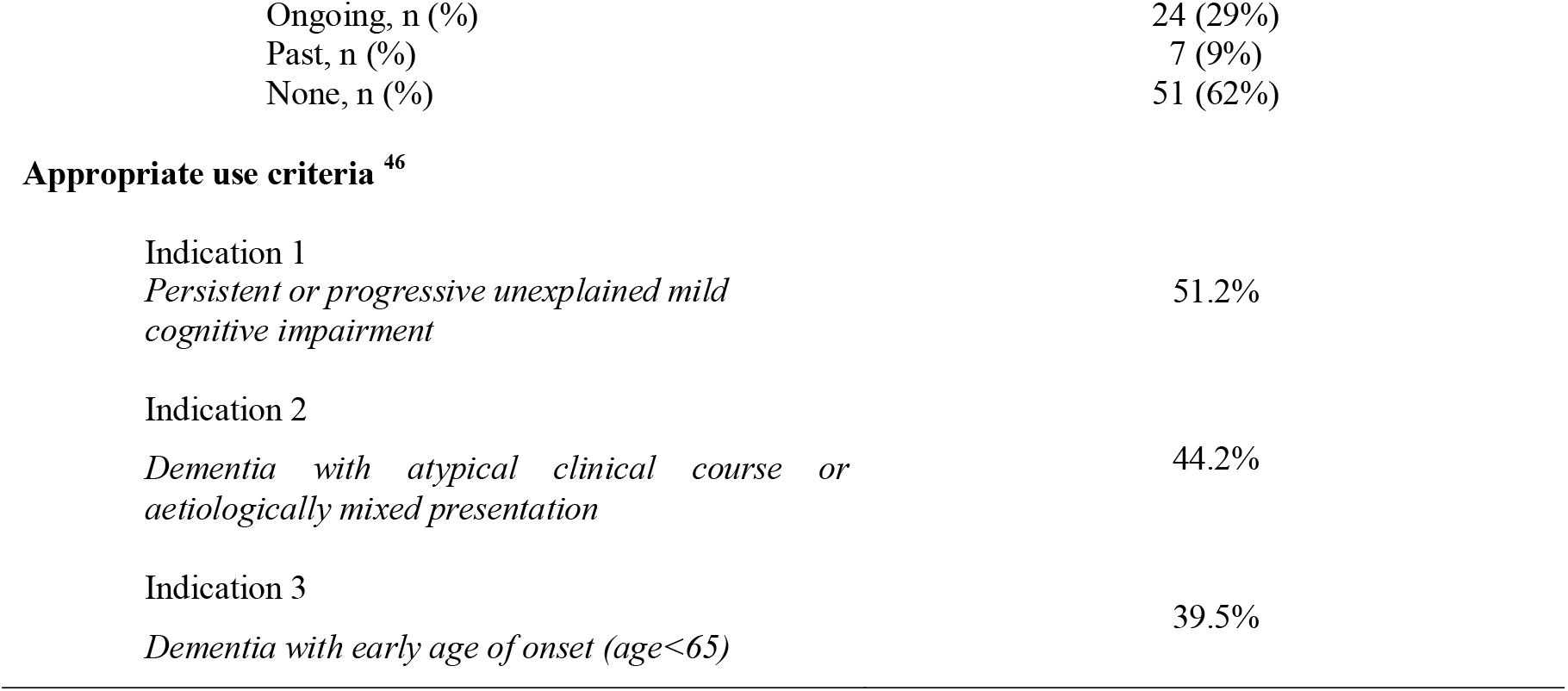
Demographic and clinical information of Alzheimer’s patients.

|  | Aβ-pos<br>(n=86) |
| --- | --- |
| **Demographics** |  |
| Mean age ± SD (years) | 67.56±8.06 |
| Age range | 49.1-87.4 |
| Sex (female), n (%) | 42 (48.8%) |
| Presentation (*Amnesic/Non-amnesic*) | 64/22 |
| Non-amnesic | 22 |
| Visuospatial, n (%) | 7 (32%) |
| Language, n (%) | 11 (50%) |
| Behavioural, n (%) | 4 (18%) |
| Alzheimer’s disease stage<br>(*Dementia/MCI*) | 48/38 |
| Depressive symptoms | 82 |
| Ongoing, n (%) | 24 (29%) |
| Past, n (%) | 7 (9%) |
| None, n (%) | 51 (62%) |
| Appropriate use criteria <sup>46</sup> |  |
| Indication 1<br>*Persistent or progressive unexplained mild cognitive impairment* | 51.2% |
| Indication 2<br>*Dementia with atypical clinical course or aetiologically mixed presentation* | 44.2% |
| Indication 3<br>*Dementia with early age of onset (age<65)* | 39.5% |

### Statistical analysis

#### Standard case-control comparisons

Cortical thickness from FreeSurfer was compared between a subgroup of patients from the *clinical cohort* (n=79) and an age- and sex-matched subgroup of healthy controls (n=79) from the ADNI *reference cohort*. Analysis of variance with age and sex as the covariates was used to compare mean overall thickness between the two groups. Region-level comparison was performed using two-tailed t-tests at each region, adjusting for multiple comparisons using the False Discovery Rate (FDR).

#### Total outlier count analysis

The *total outlier count* indicates the total number of outliers summed across all ROIs in each participant and ranges between 0 (no regions are outliers) and 148 (all regions are outliers). Outliers were defined as a z-score < -1.96. The total count of outliers does not provide information on the spatial distribution of atrophy, but it is an estimate of how widespread across the brain an individual’s deviation from the normative model is. A higher outlier count indicates significantly lower thickness compared to the reference cohort in a greater number of brain regions. The distribution of the total outlier count was tested for normality using Shapiro–Wilk test, which showed positively skewed data. Therefore, data were log-transformed to test for the effect of Group (with grouping based on sex, phenotype, disease stage, or depression history) on mean outlier count using analysis of covariance (ANCOVA), with age and sex as the covariates. Pearson’s correlation coefficient was used to test for the association between total outlier count and age at the time of MR scanning.

#### Analysis of spatial distribution of outliers

To assess the spatial distribution of cortical thickness, we computed the percentage of patients classified as outliers in each given ROI, indicating the degree of overlap in outlying regions (i.e., with lower thickness) across patients. Mapping outlier ROIs on the Destrieux atlas allowed visualization of spread and distribution at the individual- and the group-level. Analysis of variance models or Mann-Whitney non-parametric tests were run to investigate associations between clinical features and percentage of outliers in single, specified or across all ROIs. Multiple comparisons were always adjusted through Bonferroni correction. Intragroup dissimilarities in patterns of outliers were quantified using Hamming distance matrices and median Hamming distances were compared between groups (with grouping based on clinical features or disease severity). In addition, we zoomed in on the selective involvement of temporal brain regions by grouping all 30 (15 in each hemisphere) temporal gyri and sulci of the Destrieux atlas together (‘temporal’), separately from the remaining 118 (59 in each hemisphere) extratemporal gyri and sulci (‘extratemporal’) and averaging the proportion of outliers across these sets of regions. The mean percentage of ‘temporal’ outliers was compared to the mean percentage of ‘extratemporal’ outliers using an analysis of variance. To investigate whether the percentage of temporal outliers was higher in the amnestic than the non-amnestic group, a two-way ANOVA was run, testing for the interaction between outlier location and phenotype.

#### Exploratory analysis of brain-phenotype associations

Normative modelling provides the additional advantage over case-control studies of exploring the relationship between individual-level outliers and clinical features ^35^. We run three separate sets of ANCOVAs, covarying for age and sex, to investigate differences in total outlier count according to disease severity (i.e., MCI *vs*. dementia), disease phenotype (i.e., amnestic *vs* non-amnestic), or history of depression (ongoing *vs* no symptoms). When disease phenotype and depression history were used as independent variables, disease severity was included among the covariates. Total outlier count was log-transformed to meet normality assumption. Outlier maps were built to compare spatial distribution of outliers between these groups. Hamming distance matrices and median hamming distances were used to assess intra-group dissimilarity and differences in median hamming distances between group were assessed using linear regression.

#### Amyloid quantification

Here, we explored the association between global amyloid and the total outlier count as an index of neurodegeneration. Linear regression analysis was used to test for the association between total outlier count and mean SUV_R_. Outlier maps were compared between patients with higher (n=37) and lower (n=49) levels of amyloid, defined as an SUV_R_ respectively above or below the median.

### Data availability

Data not provided in the article are available upon reasonable request.

## Results

### Demographics and clinical characteristics

AD patients scanned at sites other than ICHT were excluded by the model, leaving a total of 86 patients. The mean age at the time of MRI scanning of the final group was 67.56±8.06 years (range 49.2-87.41) and 49% were females. Sample characteristics including disease stage (MCI-AD *vs* AD dementia), presenting phenotypes (amnestic *vs* non-amnestic), depression history, and amyloid PET appropriate indications met by the study sample are provided in **Table 1**.

### Clinical AD *vs*. ADNI controls cortical thickness

The two subgroups from the clinical cohort (n=79) and the ADNI controls (n=79) were matched for age (mean±SD=68.69±7.32 years and 68.71±7.23 years, respectively) and sex (%females: 50% in each group). After controlling for age and sex, mean cortical thickness was significantly lower in the patient group (mean±SD=2.29±0.13) than in the control group (mean±SD=2.46±0.11) (F_(1,154)_=88.78, p<0.001). Region-level comparisons adjusted for multiple comparisons (FDR corrected) highlighted significantly lower thickness in the AD group for 104 of the 148 (83%) analysed regions (**Supplementary Fig.2**).

### Overall total outlier count

The median number of regions classified as outliers across the clinical cohort (n=86) was 21.5 (IQR=35), with a total outlier count ranging between 1 and 120 out of the 148 ROIs. Females had significantly higher number of outliers (median=31.5, IQR=52) than males (median=17.5, IQR=33) (U=565, p=0.002), while there was no evidence of an association between age and total outlier count (r=-0.17, p=0.11).

### Regional distribution of outliers

The overall proportion of outliers was comparable between the left (median=21.5%, IQR=18%) and right (median=19%, IQR=17%) hemispheres (U=2620, p=0.65) (**Fig.1A**). Across the whole clinical cohort, the superior temporal sulcus (STS) featured the highest proportion of outliers in both the right (60%) and left (52%) hemispheres (**Fig.1B**). This means that this brain region showed significantly reduced thickness compared to the norm in just over half of AD patients. Specifically, the STS was classified as outlier in both hemispheres in 48% patients, in either the left or the right in 17% patients, and in none of the two in the remaining 35%. We further explored the characteristics of these three groups and found that patients with bi-hemispheric STS outliers had significantly lower mean cortical thickness and younger age than the other two groups (**Table 2**); moreover, they presented more often with non-amnestic symptoms and at more advanced disease stages than those with no STS outliers (**Table 2**). Bonferroni-adjusted post-hoc comparisons for the association between sex and STS status revealed a trend towards higher proportion of females in patients with left or right STS outlier than in those with no outliers in this region (p=0.057). Hamming distance matrices indicated within-group dissimilarity across the whole clinical cohort **(Fig.1, panel C**,**D**), (median= 35.25, IQR=20.75).

**Table 2.**
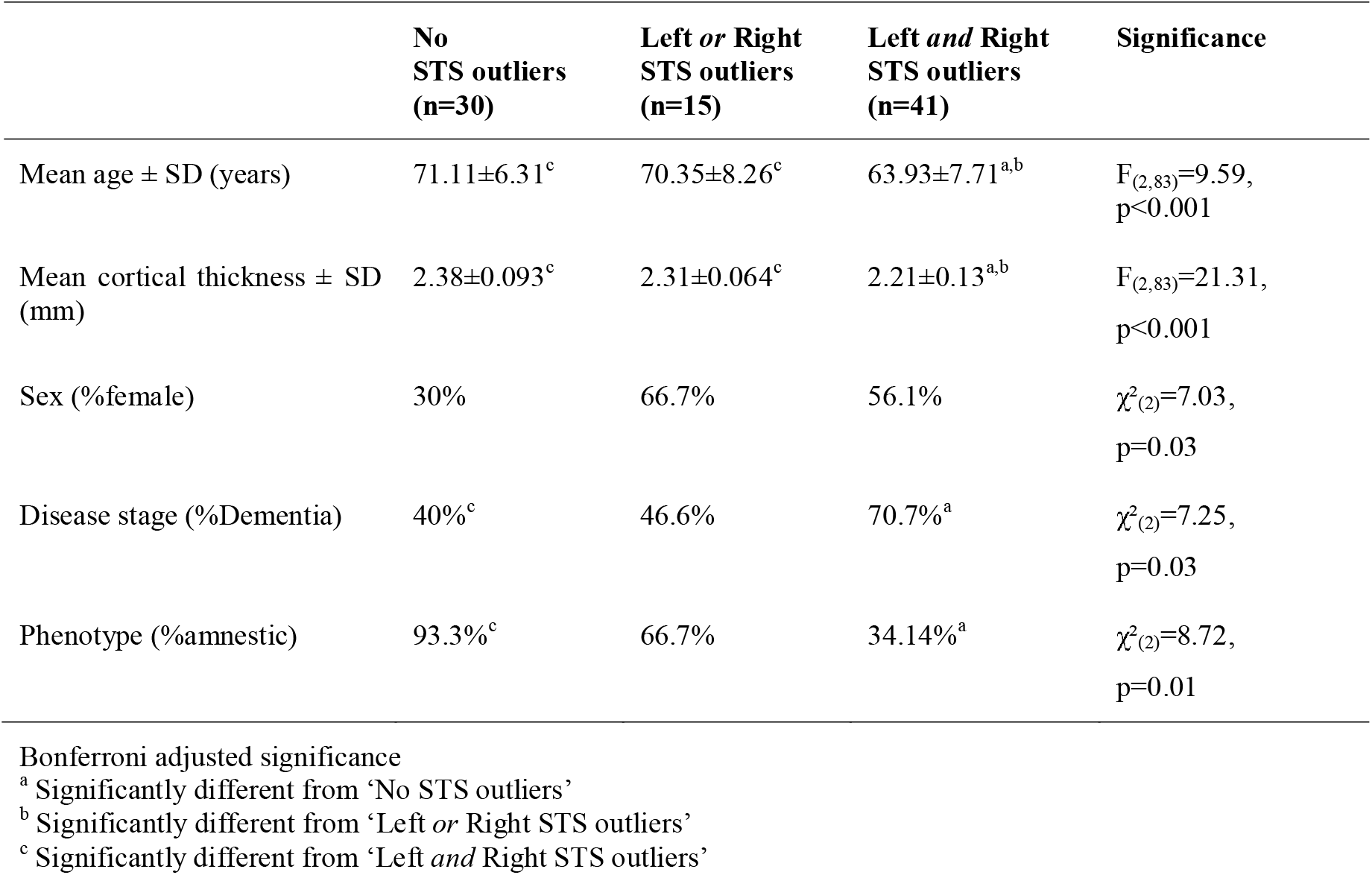
Comparison of clinical and demographic characteristics according to STS outlier status.

|  | No<br>STS outliers<br>(n=30) | Left or Right<br>STS outliers<br>(n=15) | Left and Right<br>STS outliers<br>(n=41) | Significance |
| --- | --- | --- | --- | --- |
| Mean age ± SD (years) | 71.11±6.31<sup>c</sup> | 70.35±8.26<sup>c</sup> | 63.93±7.71<sup>a,b</sup> | $F_{(2,83)}=9.59$,<br>$p<0.001$ |
| Mean cortical thickness ± SD (mm) | 2.38±0.093<sup>c</sup> | 2.31±0.064<sup>c</sup> | 2.21±0.13<sup>a,b</sup> | $F_{(2,83)}=21.31$,<br>$p<0.001$ |
| Sex (%female) | 30% | 66.7% | 56.1% | $\chi^2_{(2)}=7.03$,<br>$p=0.03$ |
| Disease stage (%Dementia) | 40%<sup>c</sup> | 46.6% | 70.7%<sup>a</sup> | $\chi^2_{(2)}=7.25$,<br>$p=0.03$ |
| Phenotype (%amnestic) | 93.3%<sup>c</sup> | 66.7% | 34.14%<sup>a</sup> | $\chi^2_{(2)}=8.72$,<br>$p=0.01$ |

Bonferroni adjusted significance
<sup>a</sup> Significantly different from 'No STS outliers'
<sup>b</sup> Significantly different from 'Left or Right STS outliers'
<sup>c</sup> Significantly different from 'Left and Right STS outliers'

**Fig. 1.**
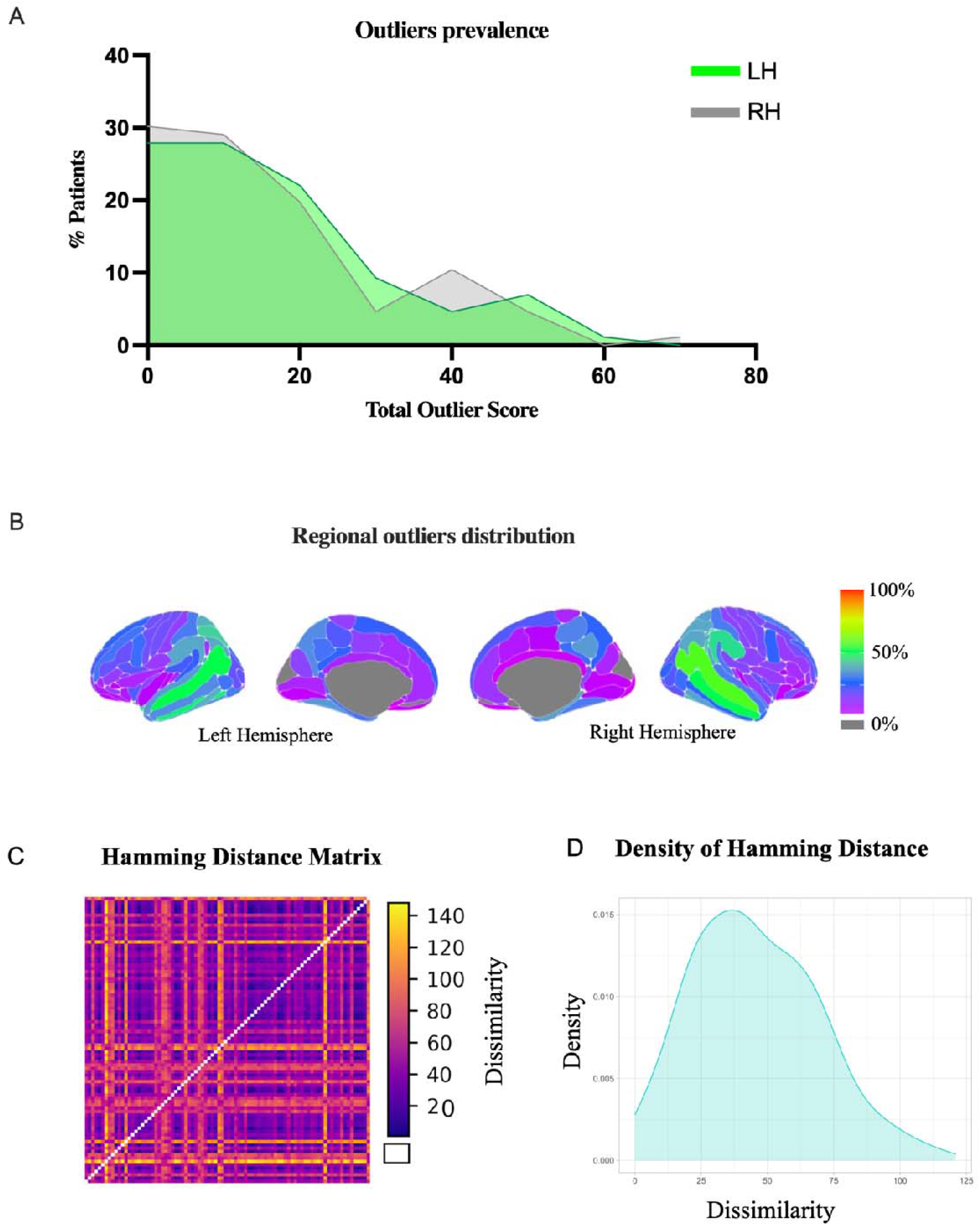
Overall outlier distribution. **(A)** Distribution of outlier prevalence across the left (LH) and right (RH) hemispheres. **(B)** Outlier maps showing spatial distribution of outliers in the clinical cohort (n=86). The superior temporal sulci (in green) featured the highest number of outliers in both hemispheres. **(C)** Hamming distance plot illustrating dissimilarity between patients in the spatial distribution of outliers. Yellow indicates greater dissimilarity. **(D)** Outlier distance density illustrates the spread of outlier dissimilarity (calculated by Hamming distance).

#### Temporal Lobe

The mean percentage of ‘temporal’ outliers was 31.5% (SD=13.7%), ranging between 7% in left lingual gyrus and 56% in the superior temporal sulcus. This was significantly higher than the extratemporal regions (F_(1,146)_=29.39, p<0.001), where the mean outlier percentage was 19.1% (SD=10.5%), ranging between 0% in the left suborbital sulcus to 47% in the right supramarginal gyrus. There was no interaction effect between outlier location (temporal *vs* extratemporal) and phenotype (amnestic *vs* non-amnestic) on the percentage of outliers (F_(1,144)_=0.003, p=0.96), suggesting comparable difference between proportion of temporal and extratemporal outliers in amnestic (mean difference=12%) and non-amnestic (mean difference=13%) patients. Notably, 12% and 20% of patients had no outliers in the left or right temporal regions respectively, and these were not more likely to be non-amnestic.

#### Disease stage

The severity of clinical impairment at the time of MR scanning was extracted from clinical records and patients were classified as MCI-AD (n=38) or AD-dementia (n=48). Analysis of covariance on log-transformed total outlier count data, using age and sex as covariates and disease severity as a predictor, revealed a significantly higher total outlier count in the AD-dementia group (median=30, range 2-120) than in the MCI-AD group (median=17.5, range 1-109) (F_(1,82)_=8.33, p=0.005). In the AD-dementia group, the highest proportion of outliers was seen at the level of the STS in both the left (67% of patients) and right (69% of patients) hemispheres. In the MCI-AD group, instead, the highest involvement of the STS was seen in the right hemisphere (50% of patients) but not in the left one where the most frequently outlying region was the planum polare (37% of patients), that is the part of the superior aspect of the superior temporal gyrus located anterior to the transverse temporal gyrus **(Fig.2A)** ^45^. Overall, in both groups there was a widespread distribution of outliers across the brain with limited overlap of outlying regions outside the temporal lobe **(Fig.2A)**. This is suggestive of high heterogeneity in the spatial patterns of atrophy which is independent of disease severity. Hamming distance matrices indicated greater within-group dissimilarity in patients within the dementia group (median=41, IQR=22), relative to the MCI group (median=28, IQR=21) **(Fig.2, panels B;C**). Linear regression revealed that participants within the dementia group were statistically more dissimilar to each other than were people in the MCI group (F_(1,73)_= 8.15, p<0.01).

**Fig. 2.**
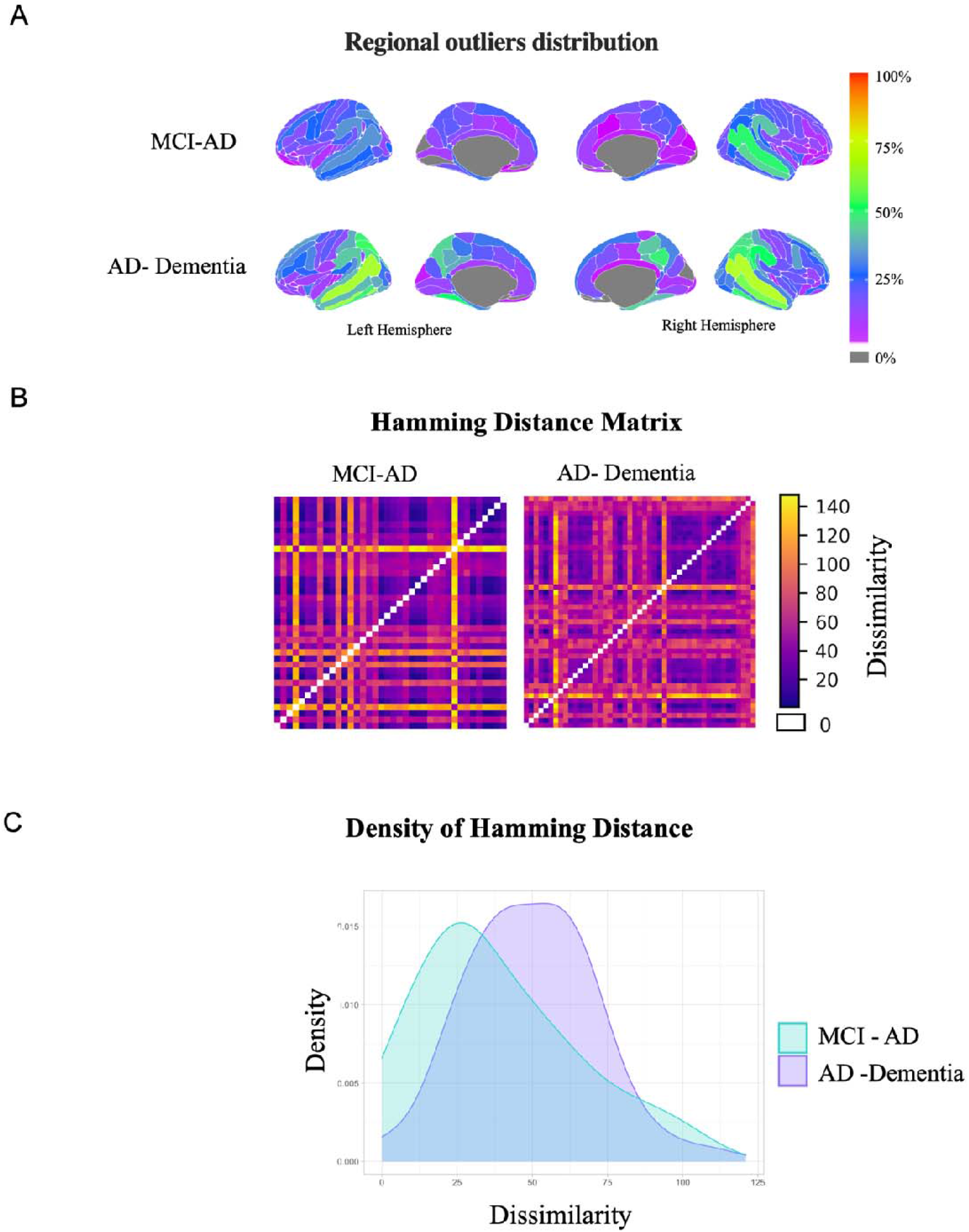
Outlier profiles according to disease severity. **(A)** Outlier maps showing distribution of outliers according to disease severity. **(B)** Hamming distance plot illustrating dissimilarity between patients in the spatial distribution of outliers; yellow indicates greater dissimilarity. **(C)** Outlier distance density illustrates the spread of outlier dissimilarity (calculated by Hamming distance).

#### Presenting phenotype

Analysis of covariance on log-transformed data, using age, sex and disease stage as covariates and phenotype as a predictor, revealed a significantly higher total outlier count in the non-amnestic (median=37.5, range 11-120) than the amnestic (median=19.5, range 1-109) group (F_(1,81)_=5.49, p=0.02). In the amnestic group, the highest proportion of outliers was seen at the level of the superior temporal sulcus in the right hemisphere (53% of patients) and in the inferior temporal sulcus in the left hemisphere (47% of patients). In the non-amnestic group, the STS was the highest involved region in both the left (77%) and right (82%) hemispheres (**Fig.3A**). Hamming distance matrices indicated greater within-group dissimilarity in patients within the non-amnestic group (median= 44.75, IQR= 15.38), relative to the amnestic group (median= 31.5, IQR= 19.62) **(Fig.3, panels B; C**). Linear regression revealed that participants within the non-amnestic group were statistically more dissimilar to each other than were people in the amnestic group (F_(1,84)_= 8.13, p < 0.01).

**Fig. 3.**
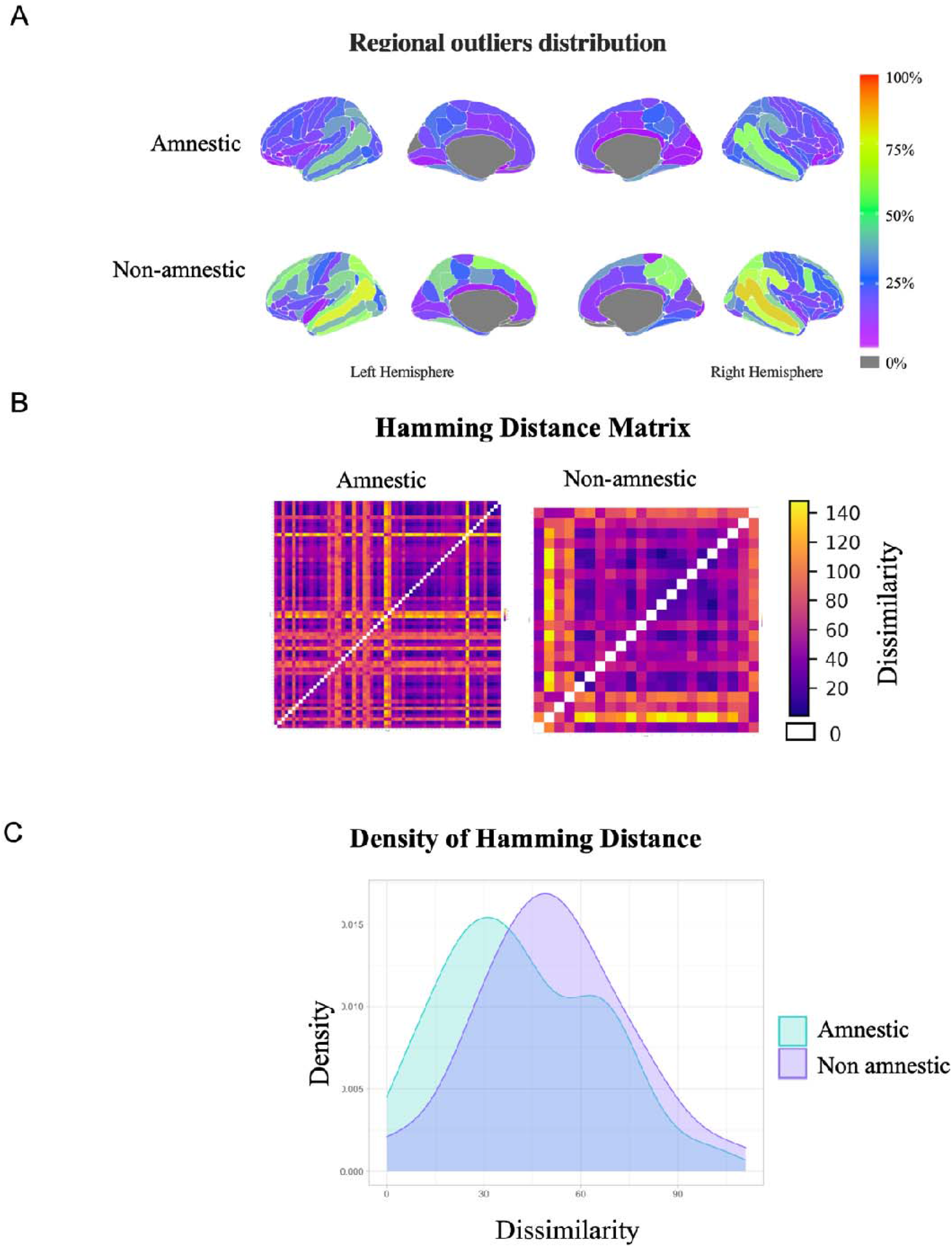
Outlier profiles according to phenotype. **(A)** Outlier maps showing distribution of outliers according to phenotype. **(B)** Hamming distance plot illustrating dissimilarity between patients in the spatial distribution of outliers; yellow indicates greater dissimilarity **(C)** Outlier distance density illustrates the spread of outlier dissimilarity (calculated by Hamming distance).

#### Comorbid depressive symptoms

Comorbid depressive symptoms are a frequent feature in patients with AD ^8^, but the nature of this link is still unclear. Where possible (n=83), we classified patients according to whether they had past (n=7), ongoing (n=24), or no (n=51) depressive symptoms at the time of presentation to our clinic (see Loreto et al., 2022 for classification procedure). After controlling for age and sex and disease severity, ANCOVA on log-transformed data showed a significantly higher total outlier count in patients without history of depression (median=30, IQR=47) than in those with ongoing depression (median=16, IQR=15) (F_(1,70)_=8.56, p=0.005). The left and right superior temporal sulci were again the regions with the highest percentage outliers in patients without (59% and 65% respectively) and with ongoing (42% and 50% respectively) depression. Other regions showing a high proportion of outliers in patients with depressive symptoms were the inferior temporal gyrus (40%), the left intraparietal sulcus (42%) and the left inferior temporal sulcus (42%) but these were also highly overlapping in those without depressive symptoms (41%, 49%, 57%, respectively). Hamming distance matrices indicated greater within-group dissimilarity in patients without depression (median= 42, IQR= 24), relative to patients with depression (median= 25.5, IQR= 8.8). Linear regression revealed that participants without depressive symptoms were statistically more dissimilar to each other than those with depressive symptoms (F_(1, 73)_= 24.69), p < 0.001).

#### Case series

**Fig. 4** provides a short case series of four patients selected from our clinical cohort who presented to our Clinic with a comparable magnitude of episodic memory impairment and spared functioning of the other cognitive domains, preserved activities of daily living, similar age of onset but different outlier maps. For example, patients *a)* and *c)* presented with similar clinical symptoms and comparable performance on cognitive screening, but regional outlier distribution in the former was limited to the left middle-anterior part of the cingulate gyrus and sulcus while it involved 67 other regions in the latter. This finding corroborates the large heterogeneity of AD atrophy profiles at presentation and indicates another possible application of normative modelling for a closer investigation of anatomo-clinical associations.

**Fig. 4.**
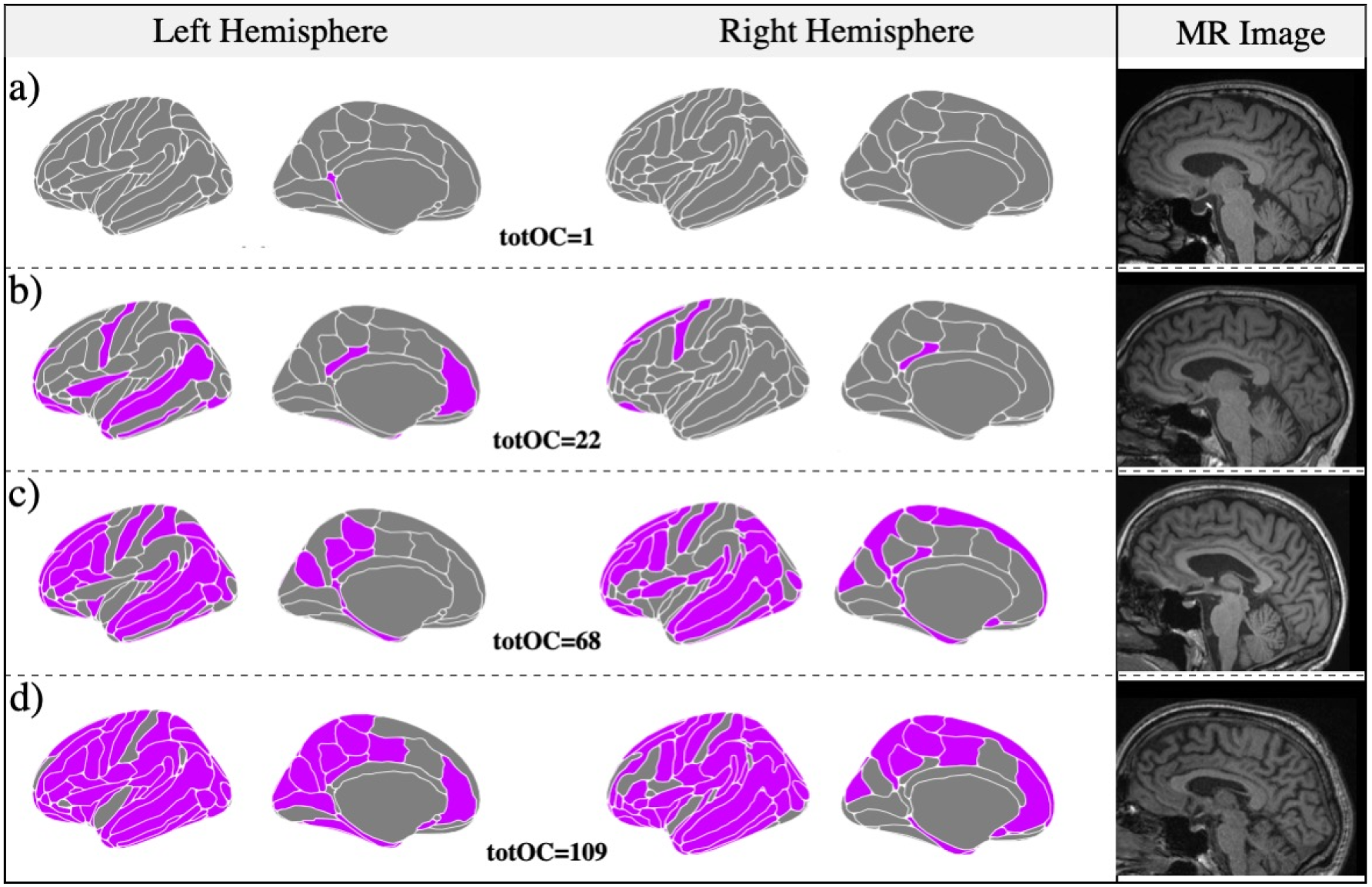
Case series. This short case series illustrates the possible use of outlier maps to gain insight into the association between atrophy profiles and clinical history. These four MCI patients had a similar clinical presentation but very heterogeneous patterns of outliers. Purple-coloured areas indicate outlier regions (z-score<-1.96). **a)** A man in his 70s presenting to our clinic with a 3-year history of memory problems, intact activities of daily living (ADLs) and preserved insight. Medical history review did not highlight significant comorbidities or presence of depressive symptoms. On examination, he scored 94/100 on the ACE-III and 26/30 on the MMSE. Clinical follow-ups revealed slow progression of cognitive deficits. **b)** A man in his late 60s presenting with a 4-year history of memory problems and preserved ADLs. Insight into the cognitive difficulties was limited and collateral account reported behavioural features such as passivity and reduced empathy. No history of depression was recorded. On examination, ACE-III score was 85/100. Follow-up visits revealed slow progression of the cognitive deficits with relative sparing of ADLs. **c)** A lady in her 70s presenting with a 2-year history of memory problems with intact ADLs, preserved insight, and no history of depression. MMSE score was 26/30. Follow-up visits revealed steady decline with gradual involvement of ADLs. **d)** A man in his mid 60s presenting with a 2-year history of memory problems and intact ADLs and no history of depression. The ACE-III score was 78/100 and follow-up highlighted clinical progression. The MMSE score at 2 years from first examination declined to 22/30. totOC: total outlier count; MCI: mild cognitive impairment; ACE-III: Addenbrooke’s Cognitive Examination version III ^61^; ADLs: activities of daily living; MMSE: mini-mental state examination ^62^

### Association between total outlier count and amyloid burden

Linear regression revealed a significant negative association between total outlier count and mean SUV_R_ (p=0.01, R^2^=0.077) (**Supplementary Fig.3A**), which survived after controlling for age (p=0.02, R^2^=0.093). A similar pattern was observed when analysing the association between mean cortical thickness and mean SUV_R_, with (p=0.03, R^2^=0.082) and without (p=0.01, R^2^=0.08) age correction (**Supplementary Fig.3B**). The lowest mean regional SUV_R_ was in the anterior division of the parahippocampal gyrus (mean SUV_R_: 1.08±0.15) while the highest was in the posterior division of the cingulate gyrus (mean SUV_R_: 1.99 ±0.31). Outlier maps and dissimilarity density plot for the low SUV_R_ and high SUV_R_ groups are provided in **Supplementary Fig.4**; notably, patients with lower levels of tracer uptake showed a higher number of outlying regions and higher within-group dissimilarity compared to those with higher SUV_R_.

## Discussion

In this study, we applied normative modelling to a real-world clinical cohort with confirmed Alzheimer’s disease to examine individual neuroanatomical variation. Patients included in this study were seen at the Imperial Memory Clinic (London, United Kingdom), referred for a structural MRI and an amyloid PET as part of their diagnostic workup, and received a biomarker-confirmed clinical diagnosis of AD ^54^. Normative modelling shifts the focus from group means and case-control comparisons to individual-level deviations from the norm, making it possible to parse spatial heterogeneity in neuroimaging profiles. Using this technique, we found that the total outlier count (i.e., count of each patient’s regions with significantly lower thickness than the normative cohort) ranged widely within patients with confirmed Alzheimer’s disease. In addition, the analysis of outlier maps (i.e., individual regional distribution of outliers, and hence of thickness, across the brain) highlighted prominent involvement of the superior temporal sulcus, but widespread distribution across the remaining regions with minimal were associated with clinical features including severity, presenting phenotype and comorbid depression.

We found that the individual magnitude of deviation from the normative model varied widely across AD patients, with the total number of outliers ranging between 1 and 120 out of 148 and a median of 21.5. This is in line with Verdi and colleagues’ previous findings on a research cohort, in which the total outlier count ranged from 0 and 106 with a median of 12, while highlighting even wider spread in the present clinical cohort ^39^. Importantly, the range of outliers was equally wide in both our MCI and dementia groups, suggesting that this finding was not attributable to disease stage. Moreover, the lack of correlation between total outlier count and age would exclude a possible confounding effect of age on the observed outlier distribution.

The examination of the spatial distribution of outliers across ROIs revealed prominent involvement of the superior temporal sulci, which were affected in up to 60% of patients, most frequently in younger and non-amnestic patients. Recent studies assessing the role of sulcal morphology in AD diagnosis have reported significantly wider superior temporal sulci in both early- and late-onset AD ^55, 56^ and negative association with memory performance ^56^. In another study, this region was also the one showing the earliest deposition of amyloid-beta in patients at risk of cognitive decline ^57^. Our findings are in line with those reported by Verdi et al.’s recent study on the ADNI cohort in which STS was among the set of temporal outlier regions differentiating AD from MCI and controls. On the other hand, in the ADNI Alzheimer’s disease group, the highest proportion of outliers was situated in the left parahippocampal gyrus and it involved 47% of patients ^39^. In the present clinical cohort, the left parahippocampal gyrus was classified as an outlier in 30% of all subjects and in 31% of the subgroup with AD-dementia. Conversely, in the ADNI cohort, the percentage of outliers in the STS ranged between 36% in the left hemisphere and 31% in the right hemisphere ^39^. Differences in the outlier maps shown in AD patients by the two studies may be due to different disease severity, phenotypic variation and, most importantly, cohort type. Alzheimer’s disease diagnosis in the ADNI study is solely based on clinical criteria and did not involve biomarker confirmation (http://adni.loni.usc.edu). Moreover, patients meeting the appropriate use criteria for amyloid PET ^43^ are a real-world cohort that, by its very nature, is more difficult to diagnose due to so-called atypical features. As such, the study of this cohort provides insight into atypical AD and its use in this clinical cohort of standard diagnostic hallmarks that originated from a disease homogeneity assumption. Notably, our analyses showed that no brain region deviated in more than 52 out of 86 clinical patients with confirmed Alzheimer’s pathology; spatial patterns of temporal lobe outliers were heterogeneous with an average overlap equal to 32%. Moreover, we identified a relatively large proportion of patients that did not significantly deviate from the norm in any of the temporal regions, despite amnestic presentation. These findings bring into question the validity of a ‘typical Alzheimer’s disease patient’.

We broadly characterised the presenting clinical picture of our cohort in terms of cognitive phenotype, disease stage, and concomitant depressive symptoms to explore anatomo-clinical associations using normative models for the first time in AD. The dementia groups showed significantly higher outlier count as well as higher dissimilarity in the regional distribution of outliers. This would support the expected greater involvement of cortical areas as disease progresses ^58^. With respect to disease phenotype, instead, the non-amnestic groups showed significantly higher total outlier count and greater within-group dissimilarity than the amnestic one. This was not surprising as the non-amnestic group would have encompassed a wider range of phenotypes, such as the visuospatial, language and dysexecutive, each with prominent involvement of different network of brain regions.

The presence of concomitant depressive symptoms was associated with a lower mean outlier count and reduced within-group dissimilarity in outlying regions. Interestingly, a recent study reported a significant association between the severity of depressive symptoms and superior temporal sulcus thickness in a group of patients with clinical AD ^59^. In our cohort, the average proportion of outliers in this region was indeed high but comparable between patients with (46%) and without (62%) depression. We did not identify any cortical regions selectively involved in patients with depression, although a different pattern may have been revealed by the analysis of subcortical structures ^60^.

The negative association between outlier count and SUV_R_ was an unexpected finding as this indicated higher cortical volumes in patients with higher amyloid pathology burden. This was further corroborated by the significant positive association between SUV_R_ and raw mean cortical thickness. It is possible that, within the group of amyloid-positive patients, the SUV_R_ starts decreasing with decreasing cortical volumes.

This study presents some limitations. First, the sample size was relatively small due to the unavailability of eligible T1-w data in clinically acquired scans. However, the very strict criteria we adopted at the time of image selection and at output evaluation ensure that the observed outliers represent clinically relevant deviations rather than deviations based on artefacts or inaccurate segmentation. Second, due to the retrospective nature of our data collection, we were not able to administer scales for granular quantification of cognitive functioning and depressive symptoms. Future studies are required to map out these relationships in addition to understanding how different pathogenic mechanisms, such as ApoE genotype, might influence the heterogeneity in the type of clinical cohort that we have described. Third, the assessment of the association between atrophy and depression was limited by the unavailability of subcortical outliers, which are currently not part of our normative model, and of information on antidepressant treatment. This highlights an important future direction in this area.

To conclude, this is the first study illustrating the possible applications of neuroanatomical normative models to parse spatial heterogeneity across individual neuroimaging data from a real-world clinical cohort of patients with confirmed Alzheimer’s disease. Our findings highlight striking variability across patients despite comparable disease stage and presentation. While case-control studies have been important in the progress of AD research, these are not well placed to advance our understanding of disease heterogeneity. As illustrated by our analysis of group differences between the AD patients and healthy control individuals, the standard case-control approach would have hidden intragroup variation and it is likely that ‘statistical outlier’ patients would have driven most of the limited case-control differences. As AD research finds its path to precision medicine, it is crucial to explore and incorporate novel methods of analysis that are as free as possible from the assumption of intragroup homogeneity. Neuroanatomical normative modelling provides a principled bridge between big data analytics and personalised medicine by shifting the analytical focus from group means to intragroup variation through the analysis of individual deviations ^15, 28^.

## Data Availability

All data produced in the present study are available upon reasonable request to the authors

## Ethics Approval

Ethical approval for this study was obtained from the Camden and Kings Cross UK Research Ethics Committee (REC number 20/LO/0442) and the Cambridge East Research Ethics Committee (REC number 10/H0304/70).

## Patient Consent for Publication

Not required.

## Acknowledgements

Data collection and sharing for this project was funded by ADNI (National Institutes of Health Grant U01 AG024904) and DOD ADNI (Department of Defence award number W81XWH-12-2-0012). ADNI is funded by the National Institute on Aging, the National Institute of Biomedical Imaging and Bioengineering, and through generous contributions from the following: AbbVie, Alzheimer’s Association; Alzheimer’s Drug Discovery Foundation; Araclon Biotech; BioClinica, Inc.; Biogen; Bristol-Myers Squibb Company; CereSpir, Inc.; Cogstate; Eisai Inc.; Elan Pharmaceuticals, Inc.; Eli Lilly and Company; EuroImmun; F. Hoffmann-La Roche Ltd and its affiliated company Genentech, Inc.; Fujirebio; GE Healthcare; IXICO Ltd.;Janssen Alzheimer Immunotherapy Research & Development, LLC.; Johnson & Johnson Pharmaceutical Research & Development LLC.; Lumosity; Lundbeck; Merck & Co., Inc.;Meso Scale Diagnostics, LLC.; NeuroRx Research; Neurotrack Technologies; Novartis Pharmaceuticals Corporation; Pfizer Inc.; Piramal Imaging; Servier; Takeda Pharmaceutical Company; and Transition Therapeutics. The Canadian Institutes of Health Research is providing funds to support ADNI clinical sites in Canada. Private sector contributions are facilitated by the Foundation for the National Institutes of Health (*www.fnih.org*). The grantee organization is the Northern California Institute for Research and Education, and the study is coordinated by the Alzheimer’s Therapeutic Research Institute at the University of Southern California. ADNI data are disseminated by the Laboratory for Neuro Imaging at the University of Southern California.

## Supplementary material

Supplementary material is available at *medrxiv* online.

