## Supplementary for "Examining real-world Alzheimer’s disease heterogeneity using neuroanatomical normative modelling"

**Supplementary Fig.1**

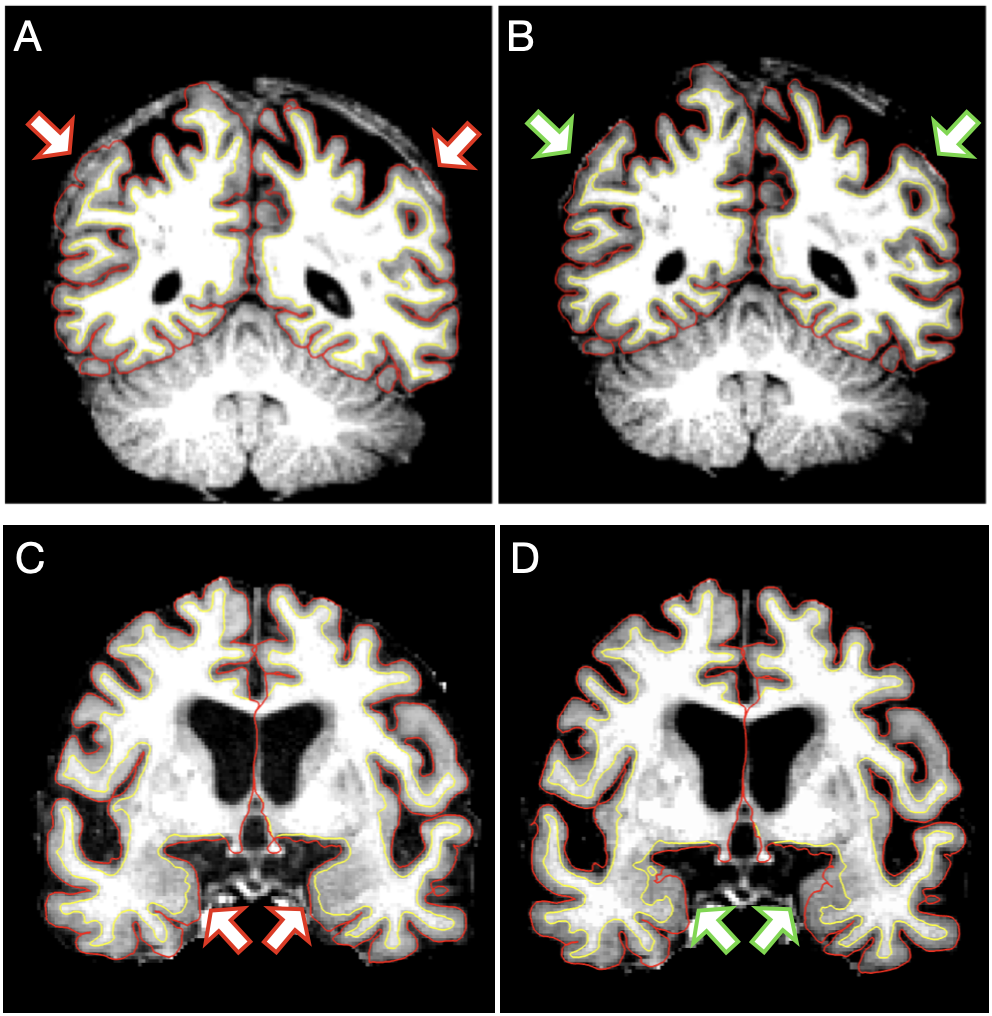

**Supplementary Fig.1** Most errors occurred in the skull stripping step (75%) leading to the erroneous inclusion of portions of dura or skull within the pial surface. This type of error was fixed by adjusting input parameters to the skull stripping step and/or by manually editing the brain mask. **(A)** Example of skull stripping error before and **(B)** after correction. Failure to identify and address this error would have led to the overestimation of parietal volumes in this subject. The remaining 25% errors occurred at the intensity normalization step (i.e., the homogenization of the signal intensity of the white and grey matter aimed at better differentiation between tissue types) leading to the exclusion of WM and/or GM voxels from the segmented surface or the inclusion of non-WM voxels in the WM surface. This type of error was fixed by adding control points to adjust voxel intensity and/or by manually adding/erasing WM voxels. **(C)** Example of intensity normalization error leading to the inclusion of non-WM voxels in the WM surface, before and **(D)** after correction. Failure to identify and address this error would have led to the overestimation of temporal lobe volumes in this subject.

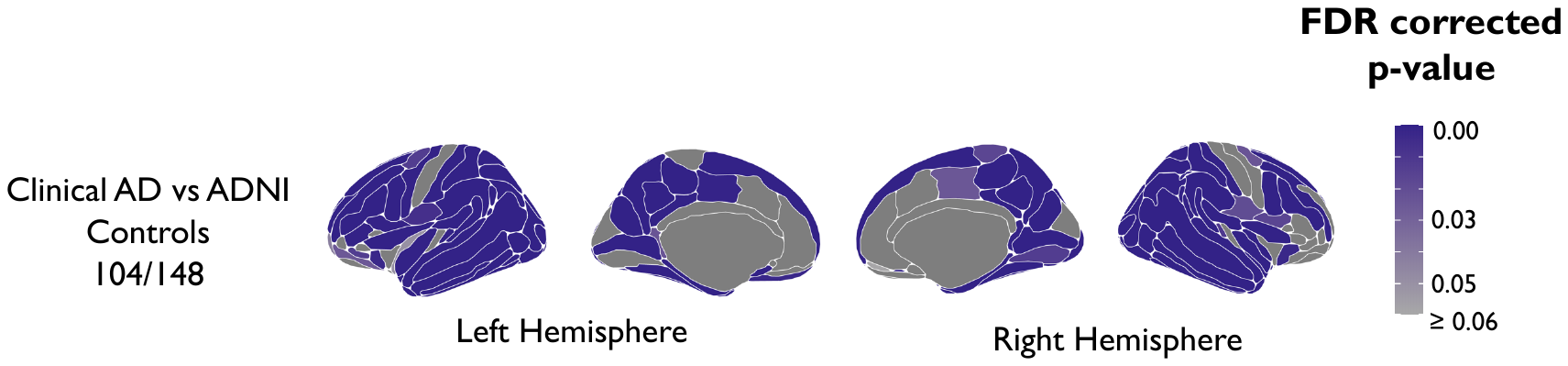

**Supplementary Fig.2.** P-value maps to illustrate significant group differences of cortical thickness at each region.

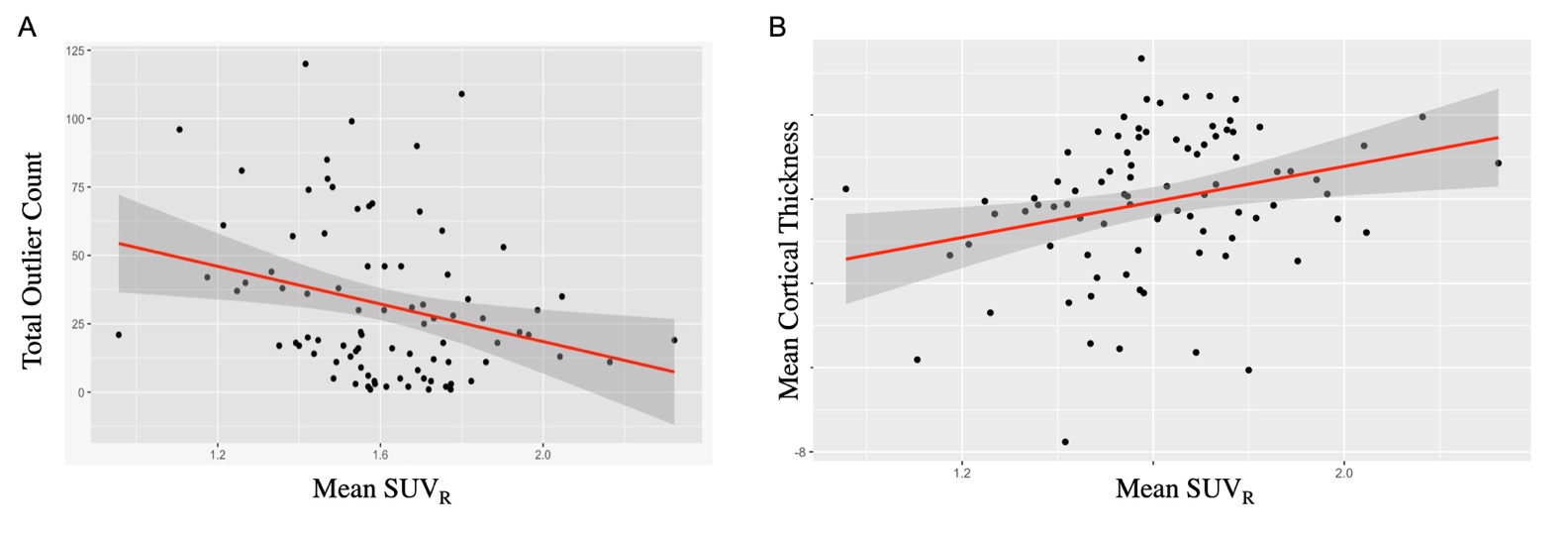

**Supplementary Fig.3** **(A)** Scatter plot showing the association between total outlier count and mean cortical SUV_R_ within the AD clinical cohort. **(B)** Scatter plot showing the association between mean cortical thickness and mean cortical SUV_R_ within the AD clinical cohort. Note: lower outlier count indicates higher cortical thickness.

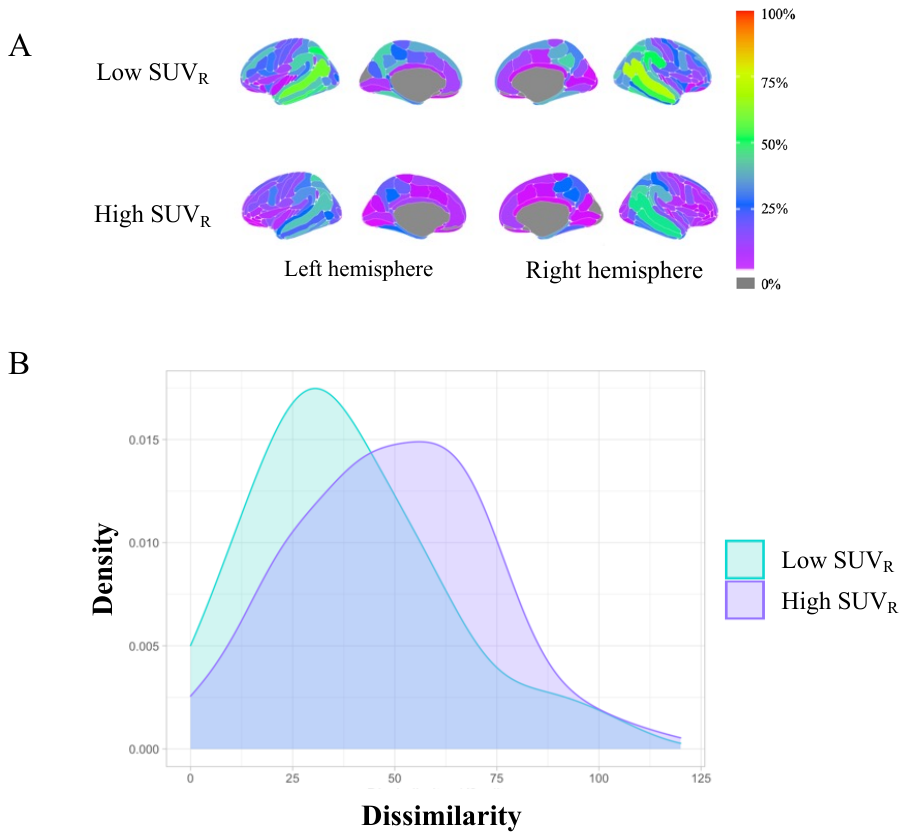

**Supplementary Fig.4** Outlier maps showing distribution of outliers according to **(A)** low or high SUV_R_. **(B)** Outlier distance density illustrates the spread of outlier dissimilarity (calculated by Hamming distance).
